# Diagnostic accuracy of age-adjusted D-dimer for pulmonary embolism among Emergency Department patients with suspected SARS-COV-2: A Canadian COVID-19 Emergency Department Rapid Response Network study

**DOI:** 10.1101/2022.03.07.22272036

**Authors:** K Lin, K Xu, R Daoust, J Taylor, R Rosychuk, JP Hau, P Davis, G Clark, A McRae, CM Hohl, the Canadian COVID-19 Emergency Department Rapid Response Network (CCEDRRN) for the NCER and the CCCTG

**Affiliations:** Department of Emergency Medicine, University of Calgary, Calgary, AB, Canada; Department of Emergency Medicine, University of British Columbia; University de Montreal, Montreal, QC; Department of Paediatrics, University of Alberta, Edmonton, AB, Canada; Department of Emergency Medicine, University of Saskatchewan, Saskatoon, SK Canada; McGill University Health Centre, Montreal, QC, Canada

## Abstract

**Importance:** Ruling out pulmonary embolism (PE) among patients presenting to the Emergency Department (ED) with suspected or confirmed SARS-COV-2 is challenging due to symptom overlap, known increased pro-thrombotic risk, and unclear D-dimer test interpretation.

**Objective:** Our primary objective was to assess the diagnostic accuracy of standard and age-adjusted D-dimer test thresholds for predicting 30-day pulmonary embolism (PE) diagnosis in patients with suspected SARS-COV-2 infection.

**Design, Setting, and Participants:** This was a retrospective observational study using data from 50 sites enrolling patients into the Canadian COVID-19 ED Rapid Response Network (CCEDRRN) registry between March 1, 2020 to July 2, 2021. Adults (**≥**18 years) with SARS-COV-2 testing performed at index ED visit were included if they had any of the following presenting complaints: chest pain, shortness of breath, hypoxia, syncope/presyncope, or hemoptysis. We excluded patients with duplicate records or no valid provincial healthcare number.

**Main Outcomes and Measures:** Our primary end point was 30-day PE diagnosis based on a positive computed tomography pulmonary angiogram (CTPA) or hospital discharge diagnosis code of PE. The outcome measure was the diagnostic accuracy of an age adjusted D-dimer strategy as compared to absolute D-dimer thresholds (500 – 5000 ng/mL).

**Results:** 52,038 patients met inclusion criteria. Age-adjusted D-dimer had a sensitivity (SN) of 96% (95% CI 93-98%) and a specificity (SP) of 48% (95% CI 48-49%) which was comparable to the most sensitive absolute threshold of 500 ng/mL (SN 98%, 95% CI 96-99%; SP 41%, 95% CI 40-42%). Other absolute D-dimer thresholds did not perform well enough for clinical reliability (SN <90%). Both age-adjusted and absolute D-dimer performed better in SARS-COV-2 negative patients as compared to SARS-COV-2 positive patients for predicting 30-day PE diagnosis (c-statistic 0.88 vs 0.80).

**Conclusions and Relevance:** In this large Canadian cohort of ED patients with suspected SARS-COV-2 infection, an age-adjusted D-dimer strategy had similar sensitivity and superior specificity to the most sensitive D-dimer threshold of 500 ng/mL for predicting 30-day PE diagnosis irrespective of SARS-COV-2 infection status. Adopting an age-adjusted D-dimer strategy in patients with suspected SARS-COV-2 may help avoid unnecessary CTPA testing without compromising safety.

**Trial Registration:** Clinicaltrials.gov, NCT04702945

**KEY POINTS:** *Question:* What is the diagnostic accuracy of age-adjusted and absolute D-dimer thresholds for investigating PE in ED patients with suspected SARS-COV-2?

*Findings:* An age-adjusted D-dimer strategy had comparable sensitivity and higher specificity for 30-day PE diagnosis compared to the most sensitive absolute threshold of 500 ng/mL irrespective of patient’s SARS-COV-2 status.

*Meaning:* Consider using an age-adjusted D-dimer threshold for PE risk stratification in ED patients with suspected SARS-COV-2.

## INTRODUCTION

Patients with SARS-COV-2 infection appear to be at elevated risk of venous thromboembolic disease (VTE), including pulmonary embolism (PE).(1) The prevalence of PE is ~5% among Canadian emergency department (ED) patients who undergo testing.(2) However, little is known about the risk of PE among ED patients with suspected SARS-COV-2 infection. Most VTE literature related to SARS-CoV-2 infection has focused primarily on the inpatient population where PE prevalence has been reported in the 13-32% range depending on illness severity,(1,3–5) but this does not fully reflect the ED population where many patients may have milder infections that do not require hospitalization.

The challenge of PE diagnosis in ED patients with suspected SARS-COV-2 is complicated by multiple factors. SARS-COV-2 infection is often not confirmed during the index ED visit, PE and SARS-COV-2 share common symptoms, and D-dimer interpretation in the setting of suspected SARS-COV-2 infection is not well understood. These factors contribute to variable practices in ED PE investigations among patients with suspected concurrent SARS-COV-2 infection and may result in overutilization of advanced diagnostic imaging such as computed tomography for pulmonary embolism (CTPA) or ventilation perfusion scanning (VQ).

Use of an age-adjusted D-dimer test threshold has been endorsed by the American College of Emergency Physicians (ACEP) as a safe and cost-effective strategy for excluding PE in low to intermediate-risk patients,(6–7) with similar sensitivity and superior specificity for diagnosing PE as compared to a standard absolute D-dimer threshold of 500 ng/mL.(6) However, the performance of age-adjusted D-dimer thresholds have not been established in ED patients with suspected SARS-COV-2 infection.

Our primary aim was to evaluate the diagnostic accuracy of an age-adjusted D-dimer strategy compared to absolute D-dimer thresholds for predicting 30-day PE diagnosis in ED patients with suspected or confirmed SARS-COV-2 infection.

## METHODS

### Study Design and Setting

This registry-based diagnostic test evaluation study used data from the multicenter Canadian SARS-COV-2 Emergency Department Rapid Response Network (CCEDRRN).(8) Specific information about CCEDRRN, including details about data collection methods, data validation processes, and a list of the 50 participating sites has been published previously. The ethics review boards of all participating sites reviewed and approved the study with a waiver for informed consent for study enrolment. This study follows the reporting recommendations outlined in the Standards for Reporting Diagnostic accuracy studies statement (STARD).(9) The funding organizations had no role in the study conduct, data analysis, manuscript preparation or submission.

### Participants

We included consecutive eligible ED visits between March 1, 2020 and July 2, 2021 that met the following criteria: 1) 18 years of age or older, 2) received SARS-COV-2OVID-19 testing in ED or within 24 hours of ED visit, and 3) presented to the ED with chest pain, shortness of breath, hypoxia, syncope, presyncope, and/or hemoptysis. This symptom list was predetermined iteratively through expert consensus and modified from prior work evaluating chief complaint descriptors with the highest yield for identifying patients likely to undergo PE investigations in a Canadian ED setting.(10) This study included patients from CCEDRRN sites that were able to demonstrate enrolment of ≥99% of eligible patients in order to minimize selection bias.

Suspected or confirmed SARS-COV-2 infection was defined as patients with symptoms meeting inclusion criteria who received a SARS-COV-2 test at or within 24 hours of index ED visit. In cases where a patient had multiple ED encounters involving SARS-COV-2 testing, the first encounter was used as the index ED visit. We excluded patient visits were excluded if they were duplicate records or if the patient lacked a valid provincial healthcare number.

### Data Collection

The CCEDRRN registry collected prespecified demographic and social variables, vital signs, symptoms, and comorbid conditions (derived from the International Severe Acute Respiratory and Emerging Infection Consortium (ISARIC) reporting form),(11) exposure risk variables, hospital laboratory and diagnostic imaging test results, SARS-COV-2 nucleic acid amplification test (NAAT) results, health resource utilization and patient outcomes. Trained research assistants abstracted data at each site using electronic medical record extraction and manual review of electronic or paper charts depending on site-specific documentation practices. Research assistants were blinded to the objectives of this analysis. Clinicians were not blinded to D-dimer test results when deciding on PE investigations. The reliability of health record data abstraction was verified by comparing key clinical variables abstracted retrospectively from the health record with prospective data collection in a sample of patients. The CCEDRRN central coordinating office conducted regular data quality checks and verified extreme and outlying values at each participating site to ensure high data quality. A national coordinator reviewed site study logs to ensure capture of ≥99% of consecutive eligible patients.

### Outcomes

The primary outcome was the diagnosis of PE at 30 days, defined as one or more of the following: CTPA positive for PE or ED or hospital diagnostic code for PE within 30 days of index ED visit.

### Sample size and precision

We targeted a 95% confidence interval of +/-2% around a point estimate of 95% for the sensitivity of D-dimer testing. Achieving this target would require a sample with 675 cases of pulmonary embolism. Assuming a PE incidence of 1.5-5% among patients tested, we estimated this would require approximately 50,000 total patients.

### Statistical Analysis

We calculated diagnostic performance of age-adjusted and prespecified absolute D-dimer test thresholds, including sensitivity (SN), specificity (SP), positive likelihood ratio (PLR), and negative likelihood ratio (NLR) with reported 95% confidence intervals. We used logistic regression to generate a receiver operator characteristics (ROC) curve with calculation of area under the curve for pre-specified absolute D-dimer thresholds (AUC, c-statistic). Age-adjusted D-dimer threshold was calculated using the standard formula of: age-adjusted threshold = 0.01 x [age-50 years]. Patients with no D-dimer values were not included in the calculation of diagnostic accuracy. Patients with no outcomes recorded were assumed not to have experienced the outcome.

The primary analysis quantified the diagnostic accuracy of age-adjusted and prespecified absolute D-dimer thresholds (500-5000 ng/mL in 500 ng/mL increments) for 30-day PE diagnosis, including sensitivity (SN) and specificity (SP) with 95% confidence intervals, as well as positive likelihood ratio (PLR) and negative likelihood ratio (NLR). We harmonized D-dimer values to standardized units (ng/mL FEU). Secondary analysis compared the diagnostic accuracy of age-adjusted and absolute D-dimer thresholds in pre-specified subgroups defined by SARS-COV-2 infection status (positive or negative).

To ensure patient privacy, a cell size restriction was enacted for cells reporting less than five counts. A p-value less than 0.05 was considered statistically significant. All analyses were conducted using R statistical software.

## RESULTS

### Trial Population

The CCEDRRN registry enrolled 125,630 patients between March 1, 2020 and July 2, 2021. A total of 52,038 patients met our inclusion criteria (Figure 1). Among included patients, 15,559 (29.9%) ultimately tested positive for SARS-COV-2. As compared to SARS-COV-2 negative patients, positive patients were slightly younger, more likely to arrive by ambulance, had fewer comorbidities, were more likely to present with shortness of breath rather than chest pain, had slightly shorter ED length of stays, and were more likely to have D-dimer ordered. Frequency of CTPA utilization, hospitalization, and 30-day PE diagnosis was similar irrespective of final SARS-COV-2 status (Table 1).

**Table 1.** Baseline characteristics of patients presenting with one or more symptoms suggestive of pulmonary embolism (according to COVID-19 status)

| Characteristic | COVID-19 Positive (N=15559) | COVID-19 Negative (N= 36479) | Cohen's Effect Size (d) |
| --- | --- | --- | --- |
| **Province** |  |  |  |
| BC | 4532 (29.1) | 9401 (25.8) |  |
| AB | 4015 (25.8) | 9649 (26.5) |  |
| SK | 245 (1.6) | 2575 (7.1) |  |
| MB | ≤5 | ≤5 |  |
| ON | 1939 (12.5) | 7774 (21.3) |  |
| QC | 4599 (29.6) | 6420 (17.6) |  |
| NB | 11 (0.1) | 180 (0.5) |  |
| NS | 218 (1.4) | 480 (1.3) |  |
| NFLD | ≤5 | ≤5 |  |
| PEI | ≤5 | ≤5 |  |
| **Demographics** |  |  |  |
| Age, median (IQR) | 55 (29.0) | 61 (34.59) |  |
| Female, N (%) | 7597 (48.8) | 18502 (50.7) |  |
| Arrival by Ambulance N (%) | 7298 (46.9) | 14004 (38.4) |  |
| **Presentation Acuity, N (%)** |  |  |  |
| CTAS 1 (Resuscitation) | 708 (4.6) | 1456 (4.0) |  |
| CTAS 2 (Emergent) | 5146 (33.1) | 14110 (38.7) |  |
| CTAS 3 (Urgent) | 8005 (51.4) | 16400 (45.0) |  |
| CTAS 4 (Less Urgent) | 1577 (10.1) | 4024 (11.0) |  |
| CTAS 5 (Non Urgent) | 96 (0.6) | 378 (1.0) |  |
| **Presenting Complaint/Symptom, N (%)** |  |  |  |
| Chest Pain | 5560 (35.7) | 15128 (41.5) |  |
| Shortness of Breath | 12663 (81.4) | 25531 (70.0) |  |
| Hypoxia* | 1050 (6.7) | 1674 (4.6) |  |
| Syncope/Presyncope | 2061 (13.2) | 6935 (19.0) |  |
| Hemoptysis | 331 (2.1) | 697 (1.9) |  |
| **Comorbidities, N (%)** |  |  |  |
| Hypertension | 5045 (32.4) | 12658 (34.7) |  |
| Diabetes | 2814 (18.1) | 5574 (15.3) |  |
| Obesity | 460 (3.0) | 769 (2.1) |  |
| CAD or CHF | 1696(10.9) | 7247 (19.9) |  |
| Pulmonary Disease □ | 2609(16.8) | 8427 (23.1) |  |
| CKD□ or Dialysis | 783 (5.0) | 2608 (7.1) |  |
| Liver Disease | 177 (1.1) | 682 (1.9) |  |
| Rheumatologic Disorder | 908 (5.8) | 3276 (9.0) |  |
| Active Cancer | 500 (3.2) | 2865 (7.9) |  |
| Tobacco Smoking or Vaping | 1506 (9.7) | 6825 (18.7) |  |
| Alcohol Misuse | ≤5 | 202 (0.6) |  |
| **Resource Utilization** |  |  |  |
| ED LOS□ in minutes, Median (IQR) | 318 (306) | 377 (347.9) |  |
| D-dimer Ordered, N (%) | 4311 (27.7) | 6526 (17.9) |  |
| CXR Ordered, N (%) | 14883 (95.7) | 34290 (94) |  |
| CTPA Ordered, N (%) | 2347 (15.1) | 6502 (17.8) |  |
| **Hospitalized, N (%)** | **6463 (41.5)** | **14919 (40.9)** |  |
| **30-day PE positive, N (%)** | **252 (1.6)** | **625 (1.7)** |  |

\*Hypoxia defined as triage text containing “hypoxia” or presenting oxygen saturation <90%. □Including asthma, pulmonary fibrosis, or other chronic lung disease. □Chronic Kidney Disease. □ED Length of Stay

**Figure 1.**
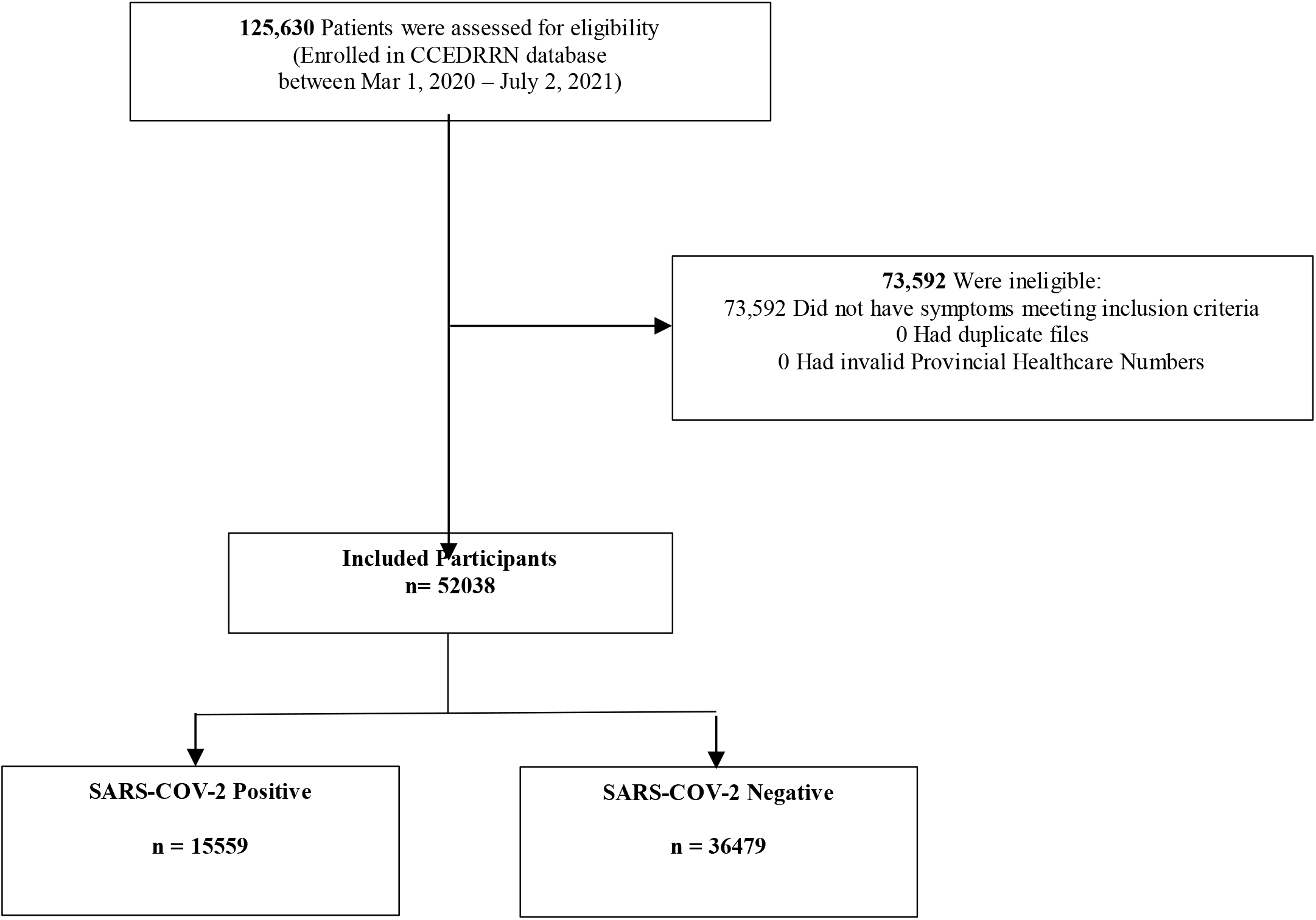
Flow Diagram of Participant Screening, Enrollment, and Cohorts According to SARS-COV-2 Status.

**Figure 2.**
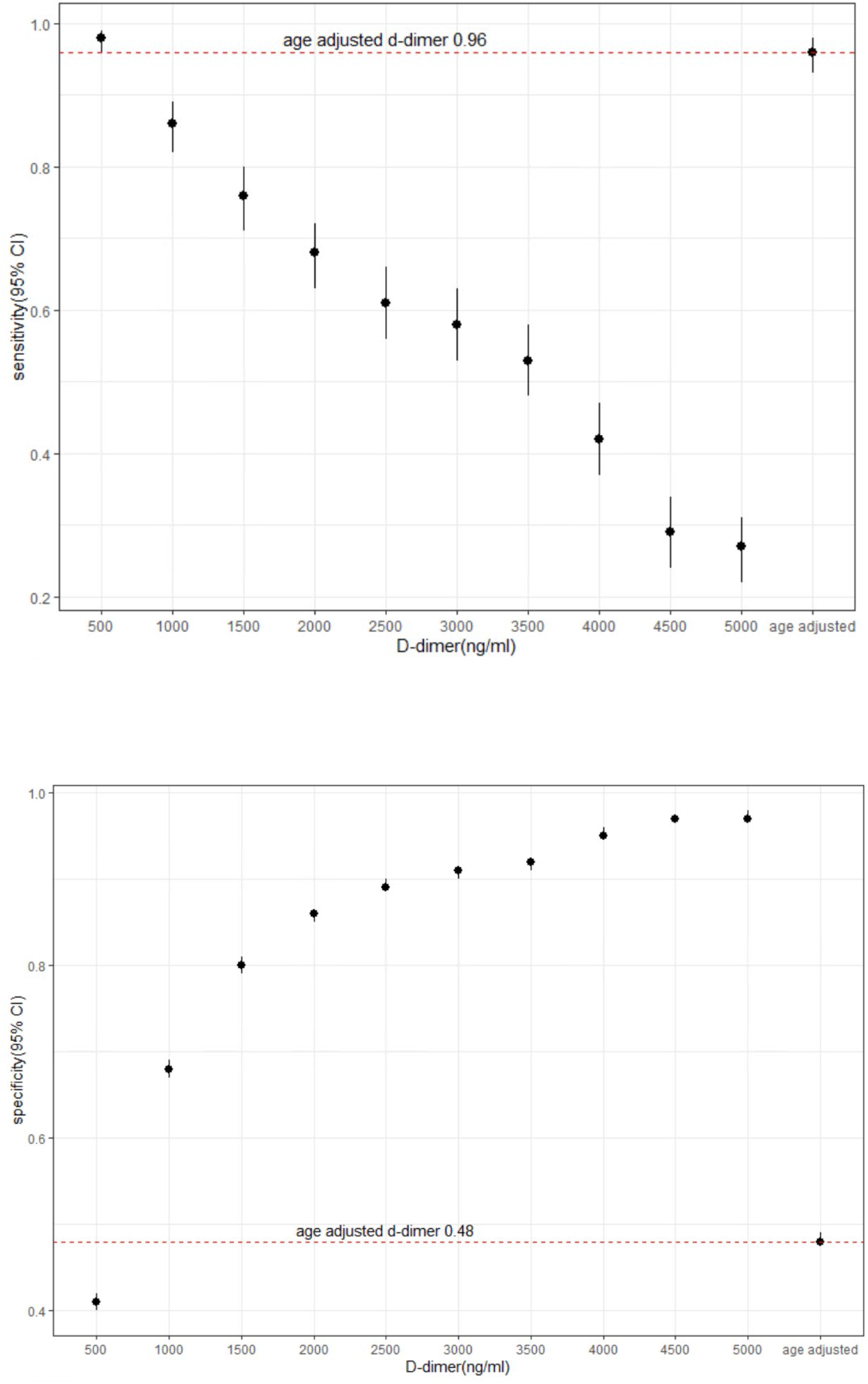
Sensitivity and Specificity of Age-adjusted vs. Absolute D-dimer Test Thresholds.

### Primary Analysis

Table 2 presents the diagnostic performance results of predefined D-dimer test positivity thresholds (including age-adjusted and absolute thresholds) for predicting 30-day PE diagnosis. Age-adjusted D-dimer had an overall sensitivity of 96% (95% CI 93-98%), specificity of 48% (95% CI 48-49%) compared to a sensitivity of 98% (95% CI 96-99%) and specificity of 41% (95% CI 40-42) of the most sensitive absolute threshold of 500 ng/mL. All other pre-specified absolute D-dimer thresholds had sensitivities of <90%. Negative likelihood ratio and positive likelihood ratio were also similar between the age-adjusted and absolute 500 ng/mL D-dimer test thresholds (NLR 0.08 vs 0.04 and PLR 1.84 vs 1.66 respectively).

**Table 2.** D-dimer Test Performance for 30-day PE Diagnosis According to Test Positivity Threshold.

| Test Positivity Threshold | 30-day PE Diagnosis |  |  |  |
| --- | --- | --- | --- | --- |
|  | Performance, % (95% CI) |  | NLR | PLR |
|  | Sensitivity | Specificity |  |  |
| 500 ng/mL | 0.98<br>(0.96 to 0.99) | 0.41<br>(0.40 to 0.42) | 0.04 | 1.66 |
| 1000 ng/mL | 0.86<br>(0.82 to 0.89) | 0.68<br>(0.67 to 0.69) | 0.20 | 2.68 |
| 1500 ng/mL | 0.76<br>(0.71 to 0.80) | 0.80<br>(0.79 to 0.81) | 0.30 | 3.80 |
| 2000 ng/mL | 0.68<br>(0.63 to 0.72) | 0.86<br>(0.85 to 0.86) | 0.37 | 4.85 |
| 2500 ng/mL | 0.61<br>(0.56 to 0.66) | 0.89<br>(0.89 to 0.90) | 0.44 | 5.55 |
| 3000 ng/mL | 0.58<br>(0.53 to 0.63) | 0.91<br>(0.90 to 0.91) | 0.46 | 6.44 |
| 3500 ng/mL | 0.53<br>(0.48 to 0.58) | 0.92<br>(0.91 to 0.92) | 0.51 | 6.62 |
| 4000 ng/mL | 0.42<br>(0.37 to 0.47) | 0.95<br>(0.95 to 0.96) | 0.61 | 8.40 |
| 4500 ng/mL | 0.29<br>(0.24 to 0.34) | 0.97<br>(0.97 to 0.97) | 0.73 | 9.67 |
| 5000 ng/mL | 0.27<br>(0.22 to 0.31) | 0.97<br>(0.97 to 0.98) | 0.75 | 9.00 |
| **Age-Adjusted D-dimer** | 0.96<br>(0.93 to 0.98) | 0.48<br>(0.48 to 0.49) | 0.08 | 1.84 |

#### Subgroup Analysis by SARS-COV-2 Status

Sensitivity was lower for most D-dimer thresholds among SARS-COV-2 positive patients as compared to SARS-COV-2 negative patients (c-statistic 0.80 vs 0.88 respectively, Table 3). Specificity among SARS-COV-2 positive patients was similar to SARS-COV-2 negative patients except for the lower specificity of the 500 ng/ml and age-adjusted thresholds, 0.31 (95% CI 0.30-0.33) vs 0.47 (95%CI 0.46-0.48) and 0.39 (95% CI 0.37-0.40) vs 0.55 (95% CI 0.54-0.56) respectively.

**Table 3.** D-dimer Diagnostic Performance and AUC (c-statistic) According to SARS-COV-2 Status.

| D-dimer Test Positivity Threshold | 30-day PE Dx |  |  |  |  |  |
| --- | --- | --- | --- | --- | --- | --- |
|  | Sensitivity (95% CI) |  |  | Specificity (95% CI) |  |  |
|  | Overall | COVID+ | COVID- | Overall | COVID+ | COVID- |
| 500 ng/mL | 0.98<br>(0.96 to 0.99) | 0.96<br>(0.91 to 0.99) | 0.99<br>(0.96 to 1.00) | 0.41<br>(0.40 to 0.42) | 0.31<br>(0.30 to 0.33) | 0.47<br>(0.46 to 0.48) |
| 1000 ng/mL | 0.86<br>(0.82 to 0.89) | 0.77<br>(0.69 to 0.84) | 0.89<br>(0.85 to 0.93) | 0.68<br>(0.67 to 0.69) | 0.64<br>(0.64 to 0.66) | 0.70<br>(0.69 to 0.71) |
| 1500 ng/mL | 0.76<br>(0.71 to 0.80) | 0.65<br>(0.56 to 0.73) | 0.79<br>(0.75 to 0.84) | 0.80<br>(0.79 to 0.81) | 0.79<br>(0.78 to 0.80) | 0.80<br>(0.79 to 0.81) |
| 2000 ng/mL | 0.68<br>(0.63 to 0.72) | 0.55<br>(0.46 to 0.64) | 0.72<br>(0.67 to 0.78) | 0.86<br>(0.85 to 0.86) | 0.86<br>(0.86 to 0.87) | 0.85<br>(0.84 to 0.86) |
| 2500 ng/mL | 0.61<br>(0.56 to 0.66) | 0.49<br>(0.40 to 0.57) | 0.66<br>(0.60 to 0.72) | 0.89<br>(0.89 to 0.90) | 0.86<br>(0.89 to 0.90) | 0.89<br>(0.88 to 0.90) |
| 3000 ng/mL | 0.58<br>(0.53 to 0.63) | 0.45<br>(0.37 to 0.54) | 0.62<br>(0.56 to 0.68) | 0.91<br>(0.90 to 0.91) | 0.91<br>(0.90 to 0.92) | 0.91<br>(0.90 to 0.91) |
| 3500 ng/mL | 0.53<br>(0.48 to 0.58) | 0.42<br>(0.34 to 0.51) | 0.56<br>(0.50 to 0.63) | 0.92<br>(0.91 to 0.92) | 0.92<br>(0.92 to 0.93) | 0.92<br>(0.91 to 0.92) |
| 4000 ng/mL | 0.42<br>(0.37 to 0.47) | 0.32<br>(0.24 to 0.40) | 0.46<br>(0.40 to 0.52) | 0.95<br>(0.95 to 0.96) | 0.96<br>(0.95 to 0.97) | 0.94<br>(0.94 to 0.95) |
| 4500 ng/mL | 0.29<br>(0.24 to 0.34) | 0.23<br>(0.16 to 0.31) | 0.30<br>(0.25 to 0.37) | 0.97<br>(0.97 to 0.97) | 0.98<br>(0.97 to 0.98) | 0.97<br>(0.96 to 0.97) |
| 5000 ng/mL | 0.27<br>(0.22 to 0.31) | 0.20<br>(0.14 to 0.28) | 0.29<br>(0.24 to 0.35) | 0.97<br>(0.97 to 0.98) | 0.98<br>(0.97 to 0.98) | 0.97<br>(0.97 to 0.97) |
| Age-Adjusted D-dimer | 0.96<br>(0.93 to 0.98) | 0.93<br>(0.88 to 0.97) | 0.97<br>(0.94 to 0.99) | 0.48<br>(0.48 to 0.49) | 0.39<br>(0.37 to 0.40) | 0.55<br>(0.54 to 0.56) |
| D-dimer C-Statistic (AUC) |  |  |  |  |  |  |
| Overall | 0.86 |  |  |  |  |  |
| COVID+ Patients | 0.80 |  |  |  |  |  |
| COVID- Patients | 0.88 |  |  |  |  |  |

## DISCUSSION

In this multicenter Canadian registry-based study involving ED patients with suspected or confirmed SARS-COV-2, an age-adjusted D-dimer test threshold had similar sensitivity and negative likelihood ratio with superior specificity to a standard 500 ng/mL absolute D-dimer test threshold. Higher absolute D-dimer test thresholds were insufficiently sensitive (<90%) to reliably rule out 30-day PE diagnosis. D-dimer sensitivity and specificity were lower in SARS-COV-2 positive patients as compared to SARS-COV-2 negative patients.

This study is unique in its ED-focused patient population. While most prior work has focused on PE evaluation in hospitalized patients with SARS-COV-2 and suggests a higher risk of PE among patients admitted for SARS-COV-2, our study captured a broad spectrum of symptomatic ED patients including nearly 60% who were discharged home from the ED. The inclusion of all suspected SARS-COV-2 patients irrespective of final confirmed SARS-COV-2 status was intended to reflect the clinical context in which most ED physicians practice, where SARS-COV-2 status is often not yet known at first point of contact when patients present with both PE and SARS-COV-2 attributable symptoms.

This study addresses the question of D-dimer performance in patients with potentially less severe illness. To our knowledge, no prior study has been able to capture such a large sample of patients. These findings suggest that an age-adjusted D-dimer strategy may safely reduce unnecessary CTPA testing without unacceptably increasing the number of missed PE cases.

## Limitations

First, certain variables of interest required for clinical VTE risk stratification rules (such as Wells and YEARS criteria) were not captured in CCEDRRN and therefore could not be validated in the study population. Second, the outcome definition of 30-day PE relied on CTPA and ED or hospital diagnostic codes which may not have completely captured all PE events. Third, the multicenter nature of this study relied on D-dimer testing across 50 ED sites with a variety of machine manufacturer models and assays utilized. All D-dimer values were harmonized to ng/mL FEU, however study results reflect an aggregate performance across all test models included and may not represent any individual test model alone. Finally, time-varying factors, such as ED crowding, prevailing SARS-COV-2 infection rates, and utilization of D-dimer as a prognostic marker of SARS-COV-2 infection severity rather than as a diagnostic tool for PE investigation were not captured in CCEDRRN and could therefore not be accounted for in the analysis.

## CONCLUSIONS

In this large Canadian cohort of ED patients with suspected SARS-COV-2 infection, an age-adjusted D-dimer strategy had similar sensitivity and negative likelihood ratio with superior specificity compared to a conservative 500 ng/mL D-dimer test threshold for predicting 30-day PE diagnosis, irrespective of SARS-COV-2 status. Adopting an age-adjusted D-dimer strategy may help avoid unnecessary CTPA testing without compromising safety in this challenging patient population.

## Supporting information

STARD Checklist

## Data Availability

CCEDRRN accepts applications for access to data by external investigators, prioritizing data requests by network Members.
https://www.ccedrrn.com/knowledge-users

## Acknowledgements

We gratefully acknowledge the assistance of Ms. Serena Small in the preparation of this manuscript. We thank the UBC clinical coordinating centre staff, the UBC legal, ethics, privacy and contract staff and the research staff at each of the participating institutions in the network outlined in the attached Supplement. The network would not exist today without the dedication of these professionals.

Thank you to all of our patient partners who shared their lived experiences and perspectives to ensure that the knowledge we co-create addresses the concerns of patients and the public. Creating the largest network of collaboration across Canadian Emergency Departments would not have been feasible without the tireless efforts of Emergency Department Chiefs, and research coordinators and research assistants at participating sites. Finally, our most humble and sincere gratitude to all of our colleagues in medicine, nursing, and the allied health professions who have been on the front lines of this pandemic from day one staffing our ambulances, Emergency Departments, ICUs and hospitals bravely facing the risks of COVID-19 to look after our fellow citizens and after one another. We dedicate this network to you.

